# Patient-Centred Communication in Lung Cancer Screening: A Clinically Focussed Evaluation of a Fine-Tuned Open-Source Model Against a Larger Frontier System

**DOI:** 10.64898/2026.04.10.26350595

**Authors:** Sanjay Khanna, Rohan Chaudhary, Navya Narula, Richard Lee

## Abstract

Lung cancer screening saves lives, yet uptake remains sub-optimal and inequitable. Personalised communication can improve attendance and reduce anxiety, but scaling such support is a workforce challenge. We fine-tuned Google’s Gemma 2 9B using QLoRA on 5,086 synthetic screening conversations and compared it against Google’s Gemini 2.5 Flash (a larger frontier model) and an unmodified baseline across 300 multi-turn conversations with 100 patient personas spanning ten clinical categories. Evaluation combined automated natural language processing metrics with independent language model judgement in two complementary modes: structured clinical rubric and simulated patient persona. The fine-tuned model achieved the highest simulated patient-experience score (3.71/5 vs 3.65 for the frontier model), recorded zero boundary violations after clinician review of all flagged instances, and led on the four most safety-critical categories. A composite Patient Adaptation Index showed that the fine-tuned model led overall (0.37 vs 0.35 vs 0.35), with its clearest advantage on the two clinically specific components: empathy calibration to patient distress and selective smoking cessation signposting. These findings suggest that targeted fine-tuning of open-source models can yield clinical communication quality comparable to larger proprietary systems, with advantages in safety-critical scenarios and suitability for NHS data governance constraints. Human clinician review of these conversations is ongoing.

## 1 Introduction

The NHS lung cancer screening programme invites individuals aged 55 to 74 with a significant smoking history for low-dose CT scanning, aiming to detect lung cancer at earlier, more treatable stages [1]. Screening has a clear survival advantage, yet uptake remains suboptimal and inequitable, with the lowest participation rates in the communities carrying the highest disease burden. At each stage, from invitation through results communication, clear and empathetic interaction can reduce anxiety and improve attendance.

Conversational AI offers one route to scaling this communication, but patients in this cohort are frequently anxious, may carry guilt about smoking history, and range widely in health literacy. Large language models have shown promise in medical question-answering [2,3], but communicative adaptation in safety-critical patient-facing settings has received less attention. Open-source models can be hosted within trust infrastructure, avoiding the data governance complexities of third-party APIs, but whether they can match a larger proprietary system’s communicative quality in a safety-critical domain has not been established.

In this work, we fine-tuned Google’s Gemma 2 9B on a synthetic dataset of lung cancer screening conversations and evaluated it alongside Google’s Gemini 2.5 Flash (hereafter: the frontier model) and the unmodified Gemma 2 9B baseline. We assessed clinical safety and communicative adaptation through automated natural language processing metrics and independent language model judgement across 300 multi-turn conversations with 100 patient personas spanning ten clinical categories relevant to the screening pathway.

## 2 Methods

### 2.1 Training Data and Fine-Tuning

We generated 5,651 synthetic multi-turn conversations using a frontier language model as teacher, guided by an NHS-aligned system prompt specifying screening programme context, scope of practice, and communication principles. Conversations were evaluated for structural integrity and clinical appropriateness, yielding 5,086 training and 565 validation examples.

We fine-tuned Google’s Gemma 2 9B Instruct using Quantised Low-Rank Adaptation (QLoRA) [4] applied to all attention and MLP projection layers, training for two epochs on a single NVIDIA L4 GPU with bfloat16 precision.

### 2.2 Evaluation Design

We constructed 100 patient personas across ten clinically motivated categories: standard consultation, anxiety, red-flag symptoms, emotional crisis, low health literacy, cultural and language considerations, scepticism, misinformation, post-screening concerns, and eligibility queries. Each persona was defined by demographic attributes, a clinical scenario, hidden concerns unknown to the chatbot, and a health literacy level (low, medium, or high).

Three model conditions were evaluated: the fine-tuned Gemma 2 9B (FT), the unmodified Gemma 2 9B Instruct (Base), and Google’s Gemini 2.5 Flash (the frontier model). Each model engaged in a multi-turn conversation with each of the 100 personas, simulated by a separate language model acting in character, producing 300 conversations totalling approximately 4,400 exchanges.

Evaluation proceeded across two tiers. Tier 1 comprised automated natural language processing metrics including boundary violation detection (instances where the model directly instructed on medication, diagnosis, or treatment rather than directing patients to professional care), readability adaptation, empathy density, cessation signposting, and a composite Patient Adaptation Index (PAI) correlating four communicative dimensions with individual patient characteristics. Tier 2 used Anthropic’s Claude as an independent judge in two framings. In the rubric mode (Tier 2a), the judge scored each conversation on seven weighted clinical dimensions. In the persona mode (Tier 2b), the judge adopted each patient’s identity and evaluated holistically whether the conversation helped, built trust, and clarified next steps. The Tier 2b score serves as the primary outcome measure.

## 3 Results

### 3.1 Patient Experience and Clinical Quality

Table 1 summarises the key evaluation outcomes.

**Table 1.** Key evaluation outcomes across three model conditions.

| Metric | FT | Base | Frontier |
| --- | --- | --- | --- |
| Tier 2b patient experience (1–5) | **3.71** | 3.24 | 3.65 |
| Patient Adaptation Index (0–1) | **0.37** | 0.35 | 0.35 |
| True boundary violations (count) | **0** | 2 | **0** |
| Red-flag referral rate | 100% | 100% | 100% |
| Adverse emotional impact | **2%** | 5% | 4% |

The fine-tuned model achieved the highest simulated patient-experience score (3.71 vs 3.65 for the frontier model), while the rubric-based composite placed it slightly below (3.88 vs 4.03). We attribute this discrepancy to residual sensitivity of structured rubrics to response length and completeness, an effect explored further in a companion abstract submitted to this conference. Clinician review of all regex-flagged conversations identified two true boundary violations, both in the baseline model: one instance of direct medication instruction during a potential cardiac event and one of unsolicited post-procedural medical advice. The fine-tuned and frontier models recorded zero true violations. Per-category analysis revealed that the fine-tuned model led on the four most safety-critical categories: red-flag symptoms (3.90 vs 3.30), emotional crisis (3.90 vs 3.70), post-screening anxiety (3.70 vs 3.10), and low health literacy (3.70 vs 3.40). The frontier model led on standard consultations (4.20 vs 3.80) and misinformation correction (4.00 vs 3.70), scenarios where comprehensive response style is beneficial.

### 3.2 Patient Adaptation

Fig. 1 presents the PAI alongside two supplementary panels that contextualise the adaptation scores. On the composite PAI (panel a), the fine-tuned model led overall (0.37 vs 0.35 for both baseline and frontier). All three conditions showed similar readability–literacy and verbosity–literacy correlations (*ρ* ≈ 0.57–0.60 and 0.32–0.38), a pattern consistent with conversational entrainment from the simulated patient input. The fine-tuned model’s advantage emerged on the two clinically specific components: empathy–distress (*ρ* = 0.37 vs 0.24 for baseline and 0.30 for the frontier model) and cessation–smoking, where it was the only condition to achieve significance (*ρ* = 0.21, *p* = 0.04), indicating selective signposting for current smokers rather than indiscriminate mention.

**Fig. 1.**
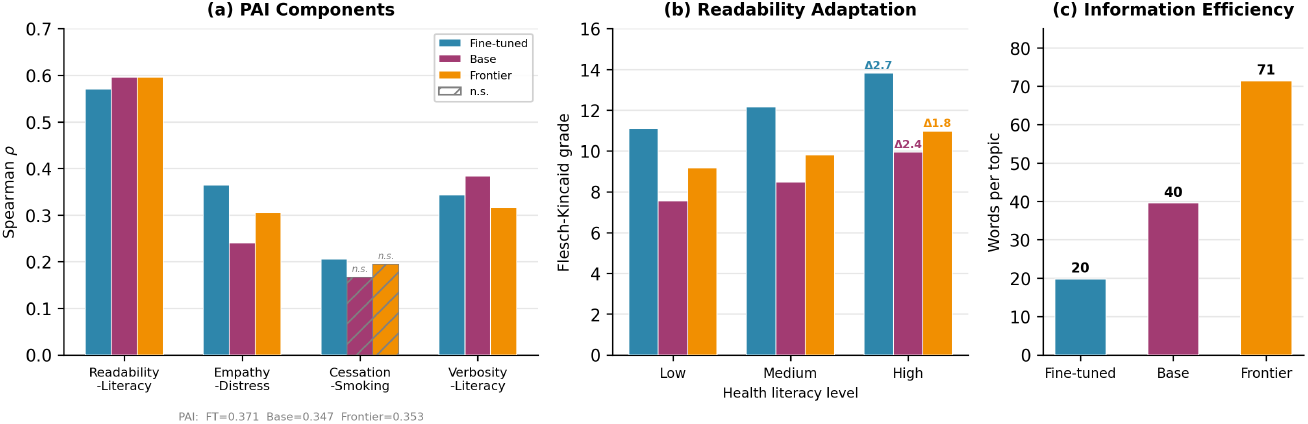
(a) Patient Adaptation Index components (Spearman *ρ*); hatched bars are non-significant (*p* ≥ 0.05). (b) Flesch–Kincaid grade by patient health literacy level; *Δ* values show the gradient between low and high literacy groups. (c) Mean words per covered clinical topic, indicating information efficiency.

Panel (b) shows that while the readability correlations were similar, the finetuned model achieved the steepest Flesch–Kincaid gradient across literacy levels (*Δ* = 2.7 FK grades between low and high literacy groups, vs 2.4 for baseline and 1.8 for the frontier model); however, the fine-tuned model’s overall reading age was higher than the frontier model’s, a limitation likely inherited from the training data that should be addressed in future iterations. Panel (c) addresses the verbosity–literacy component, where the baseline model showed a marginally higher correlation: this reflects its substantially longer responses (184 words/turn) rather than more appropriate adaptation. The fine-tuned model covered the same clinical topics in 20 words per topic, compared with 40 for baseline and 71 for the frontier model.

## 4 Discussion

These findings suggest that targeted fine-tuning of an open-source model can produce clinical communication quality that matches or exceeds a larger frontier model on the dimensions most relevant to patient-facing deployment. The fine-tuned model’s advantage is concentrated where it matters most: red-flag symptoms, emotional distress, low health literacy, and post-screening anxiety, reflecting training data that emphasised concise, contextually appropriate responses.

Gemma 2’s permissive licence and single-GPU footprint allow NHS trusts to host the model within their own infrastructure, avoiding the data governance complexities of external APIs [5]. Clinician review identified two boundary violations, both in the baseline model; the fine-tuned and frontier models recorded none.

Several limitations should be noted. All conversations used a language model as patient simulator rather than real patients, and evaluation relied on automated metrics and an independent language model judge rather than human clinician assessment. Human clinician review is ongoing and will be reported separately.

## 5 Conclusion

A fine-tuned open-source model matched a larger frontier system on simulated patient experience and surpassed it on safety-critical categories, while remaining deployable within NHS data governance constraints. Smaller models become more effective when fine-tuned on conversations appropriate to the deployment context, and evaluation of clinical communication must itself be clinically focussed, measuring not just factual coverage but the communicative qualities that determine whether patients engage with the care pathway.

## Data Availability

All data produced in the present study are available upon reasonable request to the authors

## References

1. NHS England: NHS lung cancer screening programme. NHS Long Term Plan Implementation Framework (2019)

2. Singhal, K., Azizi, S., Tu, T., et al.: Large language models encode clinical knowledge. Nature 620, 172–180 (2023)

3. Ayers, J.W., Poliak, A., Dredze, M., et al.: Comparing physician and artificial in-telligencechatbot responses to patient questions posted to a social media forum. JAMA Intern. Med. 183(6), 589–596 (2023)

4. Dettmers, T., Pagnoni, A., Holtzman, A., Zettlemoyer, L.: QLoRA: Efficient fine-tuning of quantized LLMs. In: Advances in Neural Information Processing Systems, vol. 36 (2023)

5. NHS Digital: Data security and protection toolkit. NHS England (2024)

